# Inherently explainable deep neural network-based interpretation of electrocardiograms using variational auto-encoders

**DOI:** 10.1101/2022.01.04.22268759

**Authors:** Rutger R. van de Leur, Max N. Bos, Karim Taha, Arjan Sammani, Stefan van Duijvenboden, Pier D. Lambiase, Rutger J. Hassink, Pim van der Harst, Pieter A. Doevendans, Deepak K. Gupta, René van Es

## Abstract

**Background:** Deep neural networks (DNNs) show excellent performance in interpreting electrocardiograms (ECGs), both for conventional ECG interpretation and for novel applications such as detection of reduced ejection fraction and prediction of one-year mortality. Despite these promising developments, clinical implementation is severely hampered by the lack of trustworthy techniques to explain the decisions of the algorithm to clinicians. Especially, currently employed heatmap-based methods have shown to be inaccurate.

**Methods:** We present a novel approach that is inherently explainable and uses an unsupervised variational auto-encoder (VAE) to learn the underlying factors of variation of the ECG (the FactorECG) in a database with 1.1 million ECG recordings. These factors are subsequently used in a pipeline with common and interpretable statistical methods. As the ECG factors are explainable by generating and visualizing ECGs on both the model- and individual patient-level, the pipeline becomes fully explainable. The performance of the pipeline is compared to a state-of-the-art ‘black box’ DNN in three tasks: conventional ECG interpretation with 35 diagnostic statements, detection of reduced ejection fraction and prediction of one-year mortality.

**Findings:** The VAE was able to compress the ECG into 21 generative ECG factors, which are associated with physiologically valid underlying anatomical and (patho)physiological processes. When applying the novel pipeline to the three tasks, the explainable FactorECG pipeline performed similar to state-of-the-art ‘black box’ DNNs in conventional ECG interpretation (AUROC 0·94 vs 0·96), detection of reduced ejection fraction (AUROC 0·90 vs 0·91) and prediction of one-year mortality (AUROC 0·76 vs 0·75). Contrary to state-of-the-art, our pipeline provided inherent explainability on which morphological ECG features were important for prediction or diagnosis.

**Interpretation:** Future studies should employ DNNs that are inherently explainable to facilitate clinical implementation by gaining confidence in artificial intelligence, and more importantly, making it possible to identify biased or inaccurate models.

**Funding:** This study was financed by the Netherlands Organisation for Health Research and Development (ZonMw, no. 104021004) and the Dutch Heart Foundation (no. 2019B011).

**Research into Context:** *Evidence before this study:* A comprehensive literature survey was performed for research articles on interpretable or explainable artificial intelligence (AI) for interpretation of raw electrocardiograms (ECGs) using PubMed and Google Scholar databases. Articles in English up to November 24, 2021, were included and the following key words were used: deep neural network (DNN), deep learning, convolutional neural network, artificial intelligence, electrocardiogram, ECG, explainability, explainable, interpretability, interpretable, and visualization. Many studies that used DNNs to interpret ECGs with high predictive performances were found, some focusing on tasks known to be associated with the ECG (e.g., rhythm disorders) and others identifying completely novel use cases for the ECG (e.g. reduced ejection fraction). All of these studies employed post-hoc explainability techniques, where the decisions of the ‘black box’ DNN were visualized after training, usually using heatmaps (i.e., using Grad-CAM, SHAP or LIME). In these studies, only some example ECGs were handpicked, as these heatmap-based techniques only work on single ECGs. Three studies also investigated the global features of the model by taking a summary measure of the heatmaps, by relating heatmaps to known ECG parameters (i.e., QRS duration) or by using prototypes. No studies investigated whether the features found using heatmaps were robust or reproducible.

*Added value of this study:* Currently employed post-hoc explainability techniques, usually heatmap-based, have limited explainable value as they merely indicate the temporal location of a specific feature in the individual ECG. Moreover, these techniques have been shown to be unreliable, poorly reproducible and suffer from confirmation bias. To address this gap in knowledge, we designed a DNN that is inherently explainable (i.e. explainable by design instead of investigating post-hoc). This DNN is used in a pipeline that consists of three components: (i) a generative DNN (variational auto-encoder) that learned to encode the ECG into its underlying 21 continuous factors of variation (the FactorECG), (ii) a visualization technique to provide insight into these ECG factors, and (iii) a common interpretable statistical method to perform diagnosis or prediction using the ECG factors. Model-level explainability is obtained by varying the ECG factors while generating and plotting ECGs, which allows for visualization of detailed changes in morphology, that are associated with physiologically valid underlying anatomical and (patho)physiological processes. Moreover, individual patient-level explanations are also possible, as every individual ECG has its representative set of explainable FactorECG values, of which the associations with the outcome are known. When using the explainable pipeline for interpretation of diagnostic ECG statements, detection of reduced ejection fraction and prediction of one-year mortality, it yielded predictive performances similar to state-of-the-art ‘black box’ DNNs. Contrary to the state-of-the-art, our pipeline provided inherent explainability on which ECG features were important for prediction or diagnosis. For example, ST elevation was discovered to be an important predictor for reduced ejection fraction, which is an important finding as it could limit the generalizability of the algorithm to the general population.

*Implications of all the available evidence:* A longstanding assumption was that the high-dimensional and non-linear ‘black box’ nature of the currently applied ECG-based DNNs was inevitable to gain the impressive performances shown by these algorithms on conventional and novel use cases. This study, however, shows that inherently explainable DNNs should be the future of ECG interpretation, as they allow reliable clinical interpretation of these models without performance reduction, while also broadening their applicability to detect novel features in many other (rare) diseases. The application of such methods will lead to more confidence in DNN-based ECG analysis, which will facilitate the clinical implementation of DNNs in routine clinical practice.

## Introduction

The use of deep neural networks (DNNs) has led to tremendous improvements in automated interpretation of electrocardiograms (ECGs).^1^ Recent studies have shown that DNNs achieve similar performance as cardiologists in tasks such as arrhythmia recognition and triage of ECGs.^2,3^ Even more striking, DNNs have been shown to diagnose disorders that were not yet recognized on the ECG, such as reduced ejection fraction and one-year mortality.^4,5^ Despite these promising developments, clinical implementation is severely hampered by the lack of trustworthy techniques to explain the decisions of the algorithm to clinicians.^6,7^ Due to the ‘black box’ nature of most algorithms, and the limitations of current post-hoc explainability methods, the association between input and output remains unexplainable to humans.^8^ The lack of interpretability makes it difficult for clinicians to gain enough confidence to make clinical decisions based on these algorithms, and more importantly, impossible to identify biased or inaccurate models. These issues have already been acknowledged by the new European Union’s General Data Protection Regulation, that requires a ‘right to explanation’ for AI algorithms.^9^

To improve explainability, several post-hoc explainability methods have been proposed, usually by providing heatmaps on top of the ECG. However, a major limitation of these methods is that they only provide the temporal location of ECG features important in making the diagnosis, but do not indicate the actual feature (e.g. when the QRS-complex is highlighted the feature could be R-wave height, QRS shape or something else).^5,10,11^ This makes heatmaps susceptible to confirmation bias, as we assume that the feature we think is important is also the one that was used.^6^ Therefore, instead of explaining the ‘black box’ after it was trained, the preferred way for algorithms to produce trustworthy explanations is to develop models that are inherently explainable (i.e. explainable by design).^8^

We hypothesized that an ECG can be explained by a few underlying anatomical and (patho)physiological factors of variation. Variational auto-encoders (VAE) are generative networks that use the power of DNNs to learn to compress any ECG into a selected number of explanatory and independent factors. Moreover, they can reconstruct the ECG from these factors.^12,13^ In this study, we aimed to use a VAE to identify the underlying factors of variation in the ECG and use them to develop a fully end-to-end explainable pipeline for the interpretation of ECGs. Firstly, we investigate the learned factors by relating them to known ECG parameters and the most common conventional diagnostic ECG statements. Secondly, we train and validate the explainable pipeline for use in the novel ECG use cases, detection of reduced ejection fraction and prediction of one-year mortality, and perform a comparison with current state-of-the-art ‘black box’ DNNs and conventional ECG algorithms.

## Methods

### Study participants

The dataset consisted of all patients between 18 and 85 years of age with at least one ECG acquired in the University Medical Center Utrecht (UMCU) between July 1991 and August 2020. All data were de-identified in accordance with the EU General Data Protection Regulation and written informed consent was not required by the UMCU ethical committee.

### Data acquisition for training and validation of the VAE model

All resting 12-lead ECGs were exported from the MUSE ECG system (MUSE version 8; GE Healthcare, Chicago, IL, USA) in raw voltage format and converted to median beats as described before.^10^ All ECGs that were deemed technically inadequate by either the MUSE 12SL algorithm or interpreting physician were excluded from the analyses. No labels were used in the training of the unsupervised auto-encoder.

### Data acquisition for training and validation of the ‘black box’ DNNs and explainable FactorECG pipelines

For training of the algorithms to detect conventional diagnostic ECG statements, we included a subset of ECGs that were obtained at all non-cardiology departments, as these ECGs were systematically annotated by a physician as part of the regular clinical workflow. We selected the 35 most common diagnostic statements for training (an overview can be found in the Supplementary Methods) and used 20% of the patients for hyperparameter optimization. For validation of the ECG interpretation models, an independent dataset comprising 1000 randomly selected ECGs of unique patients was annotated by a panel of 5 practicing electrophysiologists or cardiologists for all diagnostic statements as described before.^3^ A reduced set of the 35 diagnostic statements was tested, as some abnormalities did not occur in the test dataset. Moreover, the myocardial ischemia labels in different locations were combined.

To train and validate the algorithms to detect reduced ejection fraction (below 35%) and predict one-year mortality, we selected patients using the same approaches as Attia et al. and Raghunath et al., respectively.^4,5^ For the reduced ejection fraction model, patients with an ECG-echocardiogram pair (acquired within 14 days) were retrieved, the ejection fraction (EF) was dichotomized at 35% and patients were split in a 75:25 manner on the patient level. For the one-year mortality model, all patients with at least one year of follow-up available for evaluation of all-cause mortality were selected and split in a 60:40 manner on the patient level. Detailed information on the data acquisition for all three tasks can be found in the Supplementary Methods.

### VAE model architecture and training

The VAE consists of three parts: the encoder, the latent space (with multiple continuous factors, referred to as the FactorECG) and the decoder.^12^ The original 12-lead median beat ECG is entered into the encoder, that compresses the ECG to its FactorECG with 32 continuous factors. From those same factors, the ECG is reconstructed by the decoder, and the difference between the input and reconstructed ECG was used to train the model. The decoder and encoder are a standard convolutional neural network and the inverse of that neural network, respectively. A specific type of variational autoencoder was used, called the β-VAE, where an additional hyperparameter β is included in the loss term to learn disentangled factors, i.e. generative factors of variation that are independent of each other.^13^ A schematic overview of the technique can be found in Figure 1, while an animation of the approach is included as Supplementary Material. Detailed information on the training and architecture of the VAE can be found in the Supplementary Methods.

**Figure 1.**
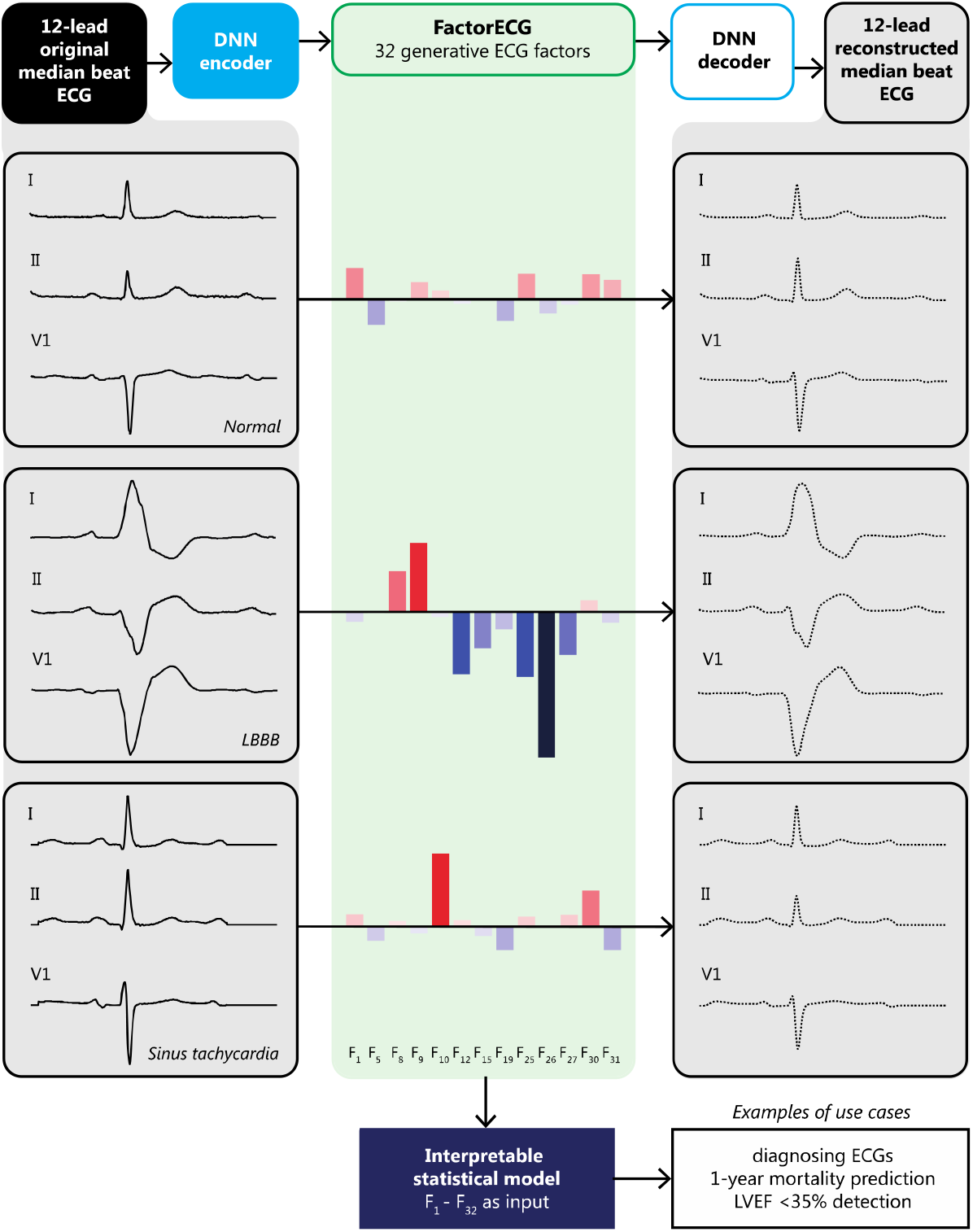
Illustration of the full pipeline: a variational auto-encoder, the FactorECG and reconstructions. The variational auto-encoder (VAE) consists of three parts: the encoder, the FactorECG space and the decoder. An input 12-lead median beat ECG is entered into the decoder, that compresses the ECG to its FactorECG with 32 continuous factors. From those same factors, the ECG is reconstructed and the difference between the input and reconstructed ECG is used to train the model. The ECG factors are subsequently used in two ways: for development of interpretable classifiers for ECG diagnostic statements, reduced ejection fraction and one-year mortality, and for visualization purposes. ECG factors can provide both individual patient- and model-level visualizations. Individual visualizations are depicted here, where three median beat ECGs and their reconstructions are represented in the FactorECG. Notably, as dimension 10 encodes ventricular frequency, we see high values for the sinus tachycardia ECG. Moreover, as dimension 26 inversely encodes left bundle branch conduction delay, we see low values for the left bundle branch block ECG. The normal ECG has value around zero for all factors, as the VAE is forced to learn factors with zero mean. ECG: electrocardiogram, LVEF: left ventricular ejection fraction, LBBB: left bundle branch block.

### VAE model explainability

The VAE model is explainable on both the model- and individual patient level. On the model-level, ECG factors are visualized by factor traversals: varying the values of an individual factor while reconstructing and plotting the median ECG beat. Every visualization starts with zeros for all factors, which represents the mean ECG in the training dataset. Then, for every individual factor, values between −5 and 5 are assigned, while keeping the others at zero, and a new generated ECG is obtained. These reconstructions are subsequently visualized in the same graph. This allows for detailed visualizations of morphological changes. On the individual patient-level, explainability is obtained by combining the distinct FactorECG values of that ECG with knowledge on the predictors that were important for a specific task. For example, if an ECG has a high value for a specific ECG factor and this factor was associated with the outcome, this would explain why this specific ECG has a higher risk of the outcome.

### Training of the baseline ‘black box’ models and explainable FactorECG pipeline

The explainable FactorECG pipeline is compared to current state-of-the-art ‘black box’ DNNs in three tasks: conventional ECG interpretation, detection of reduced ejection fraction and prediction of one-year mortality. Firstly, for all three tasks investigated, a baseline state-of-the-art ‘black box’ DNN with a similar architecture as the encoder of the VAE and the median beat ECG as input was trained.10,14 Secondly, to train the explainable FactorECG pipeline, median beat ECGs were encoded in their FactorECG, and the FactorECG values were entered into common interpretable statistical models. For the conventional ECG interpretation task, we trained binary logistic regression models for each of the 35 diagnostic ECG statements on the FactorECGs, as it provided maximum interpretability. For the detection of reduced ejection fraction and prediction of one-year mortality, as the aim was maximum performance, we trained two extreme gradient boosting decision tree (XGBoost) models.^15^ For this model, interpretability was obtained using Shapley Additive exPlanations (SHAP), which can provide feature importance measures for every ECG factor on a model- and individual patient-level.^16^ Additional information on the baseline model and training procedures for the three tasks are available in the Supplementary Methods.

### Statistical analysis

All data are presented as mean ± SD or median with interquartile range, where appropriate. All individual ECG factors were related to the conventional ECG features computed by the MUSE algorithm (i.e., ventricular rate, PR, QRS and Bazett corrected QT duration, and R and T axis) using hexagon plots and Pearson correlation coefficients. Discriminatory performance of the models is assessed in the test sets using the c-statistic or area under the receiver operating curve (AUROC) and the area under the precision-recall curve (AUPRC). As all models are weighted for class imbalance, a probability cut-off of 50% was used. Overall, 95% confidence intervals are obtained using 2000 bootstrap samples. The Transparent Reporting of a Multivariable Prediction Model for Individual Prognosis or Diagnosis Statement for the reporting of diagnostic models was followed.^17^

## Results

### Development of the VAE and explainability of the FactorECG

The dataset for training of the VAE consisted of 1,144,331 12-lead median beat ECGs of 251,473 unique patients. The VAE was able to reconstruct the median beat ECGs excellently with a mean Pearson correlation coefficient of 0·90 (p < 0·001) between the original and reconstructed ECG. Reconstructions were most accurate for sinus rhythm, sinus bradycardia, early repolarization, and pericarditis ECGs (mean r=0·91–0·92), and least accurate for the rarer ECGs with ST elevation suspected of myocardial infarction and ventricular tachycardia (mean r=0·62–0·70). An overview of mean correlation coefficients per diagnostic ECG statements can be found in Supplementary Table 1. By analyzing the factor traversals (Supplementary Figure 2), only 21 of the 32 factors were found to be necessary to reconstruct the ECG, and the other 11 were not used by the model to encode significant data. Model-level explainability, using factor traversals, is shown for a subset of the 21 factors in Figure 2. An online tool to visualize the generated ECGs interactively is available via https://decoder.ecgx.ai. To further investigate and gain interpretability in the ECG factors, Pearson correlation coefficients were computed between conventional ECG parameters and ECG factors values (Figure 3). Ventricular rate is mostly correlated to factor 10 (r=0·96, p<0·001), while QRS duration is mostly correlated to factor 25 (r=-0·47, p<0·001). PR and QT interval are mostly correlated to factors 8 (r=0·62, p<0·001) and 30 (r=-0·52, p<0·001), respectively.

**Table 1.** Diagnostic performance values for the conventional ECG interpretation task in the expert-annotated test set. The AUROC and AUPRC scores per diagnostic statement in the ECG interpretation task for the rule-based MUSE algorithm, explainable DNN model, and ‘black-box’ DNN are shown. A reduced set of the 35 diagnostic statements was tested, as some abnormalities did not occur in the test dataset. Moreover, the myocardial ischemia labels in different locations were combined. AUROC: area under the receiver operating curve, AUPRC: area under the precision-recall curve, AV: atrioventricular, CI: confidence interval, DNN: Deep Neural Network, LAFB: left anterior fascicular block, LBBB: left bundle branch block, NICD: non-specific intraventricular conduction delay, RBBB: right bundle branch block.

| Diagnostic statement | Prevalence | MUSE 12 SL |  | Explainable DNN |  | Black box DNN |  |
| --- | --- | --- | --- | --- | --- | --- | --- |
|  | n (%) | AUROC [95% CI] | AUPRC | AUROC [95% CI] | AUPRC | AUROC [95% CI] | AUPRC |
| Sinus rhythm | 697 (72) | 0.90 [0.88 - 0.92] | 0.96 | 0.94 [0.92 - 0.96] | 0.96 | 0.96 [0.95 - 0.97] | 0.98 |
| Sinus bradycardia | 30 (3.1) | 0.70 [0.61 - 0.78] | 0.09 | 0.95 [0.92 - 0.98] | 0.39 | 0.94 [0.87 - 0.97] | 0.37 |
| Sinus tachycardia | 91 (9.4) | 0.95 [0.92 - 0.97] | 0.75 | 0.99 [0.98 - 0.99] | 0.81 | 0.99 [0.99 - 1.00] | 0.94 |
| Atrial fibrillation | 90 (9.3) | 0.88 [0.84 - 0.93] | 0.73 | 0.99 [0.98 - 0.99] | 0.78 | 0.98 [0.97 - 0.99] | 0.86 |
| Atrial flutter | 2 (0.2) | 0.74 [0.49 - 1] | 0.04 | 0.98 [0.96 - 0.99] | 0.04 | 1.00 [0.99 - 1.00] | 0.67 |
| Supraventricular tachycardia | 18 (1.9) | 0.58 [0.5 - 0.67] | 0.15 | 0.97 [0.95 - 0.98] | 0.33 | 0.98 [0.96 - 0.99] | 0.34 |
| Junctional bradycardia | 4 (0.4) | 0.75 [0.5 - 1] | 0.13 | 0.99 [0.96 - 1.00] | 0.46 | 1.00 [0.99 - 1.00] | 0.56 |
| Ventricular tachycardia | 2 (0.2) | 0.50 [0.5 - 0.5] | 0 | 0.99 [0.98 - 1.00] | 0.21 | 1.00 [0.99 - 1.00] | 0.23 |
| Pacemaker rhythm | 27 (2.8) | 0.92 [0.85 - 0.98] | 0.74 | 0.97 [0.94 - 0.98] | 0.46 | 0.97 [0.93 - 0.99] | 0.68 |
| First degree AV block | 57 (5.9) | 0.86 [0.8 - 0.92] | 0.66 | 0.98 [0.97 - 0.99] | 0.68 | 0.96 [0.94 - 0.98] | 0.71 |
| Third degree AV block | 1 (0.1) | 0.5 [0.5 - 0.5] | 0 | 1.00 [1.00 - 1.00] | 0.31 | 1.00 [0.99 - 1.00] | 0.14 |
| RBBB | 59 (6.1) | 0.95 [0.91 - 0.98] | 0.66 | 0.98 [0.97 - 0.99] | 0.69 | 0.99 [0.98 - 1.00] | 0.83 |
| LBBB | 22 (2.3) | 0.88 [0.79 - 0.97] | 0.64 | 1.00 [0.99 - 1.00] | 0.82 | 1.00 [1.00 - 1.00] | 0.95 |
| LAFB | 71 (2.4) | 0.64 [0.59 - 0.69] | 0.29 | 0.84 [0.79 - 0.88] | 0.28 | 0.97 [0.96 - 0.98] | 0.62 |
| NICD | 14 (1.5) | 0.63 [0.53 - 0.76] | 0.09 | 0.94 [0.92 - 0.96] | 0.12 | 0.88 [0.73 - 0.97] | 0.3 |
| Myocardial infarction | 66 (6.8) | 0.6 [0.55 - 0.65] | 0.19 | 0.77 [0.72 - 0.82] | 0.16 | 0.77 [0.71 - 0.82] | 0.19 |
| Left ventricular hypertrophy | 44 (4.6) | 0.79 [0.71 - 0.86] | 0.32 | 0.82 [0.77 - 0.87] | 0.15 | 0.97 [0.95 - 0.98] | 0.63 |
| Low QRS voltage | 40 (4.2) | 0.76 [0.68 - 0.83] | 0.36 | 0.8 [0.74 - 0.86] | 0.18 | 0.96 [0.94 - 0.98] | 0.63 |
| Prolonged QT interval | 22 (2.3) | 0.69 [0.6 - 0.8] | 0.14 | 0.95 [0.91 - 0.97] | 0.43 | 0.93 [0.89 - 0.95] | 0.2 |
| Early repolarisation | 23 (2.4) | 0.52 [0.5 - 0.57] | 0.04 | 0.96 [0.93 - 0.98] | 0.45 | 0.98 [0.97 - 0.99] | 0.61 |
| Acute pericarditis | 7 (0.7) | 0.57 [0.5 - 0.71] | 0.15 | 0.99 [0.99 - 1.00] | 0.49 | 0.99 [0.96 - 1.00] | 0.61 |

**Figure 2.**
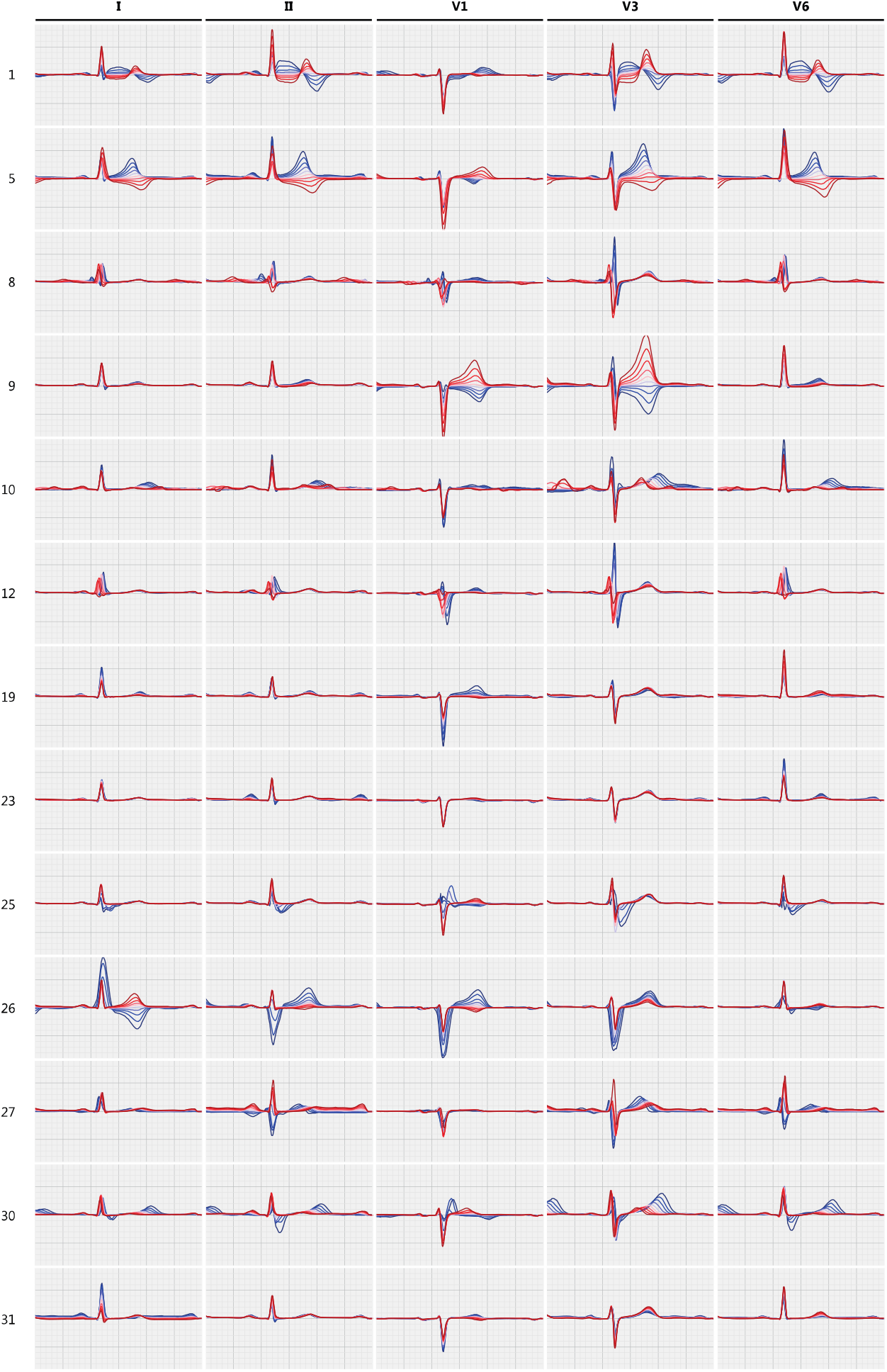
Factor traversals of a subset of the ECG factors for leads I, II, V1, V3, V6. Factor traversals of a subset of the 21 ECG factors that hold significant information for correctly reconstructing ECGs. Each row corresponds to the factor traversal for one ECG factor and the columns to a subset of the 12 leads. The factor traversal for one row is obtained by starting with a ‘mean’ FactorECG where all factors are zero and adding offsets for that factor in a range of −5 to 5. The generated ECGs are then plotted where red represents high values for that factor and blue low values.

**Figure 3.**
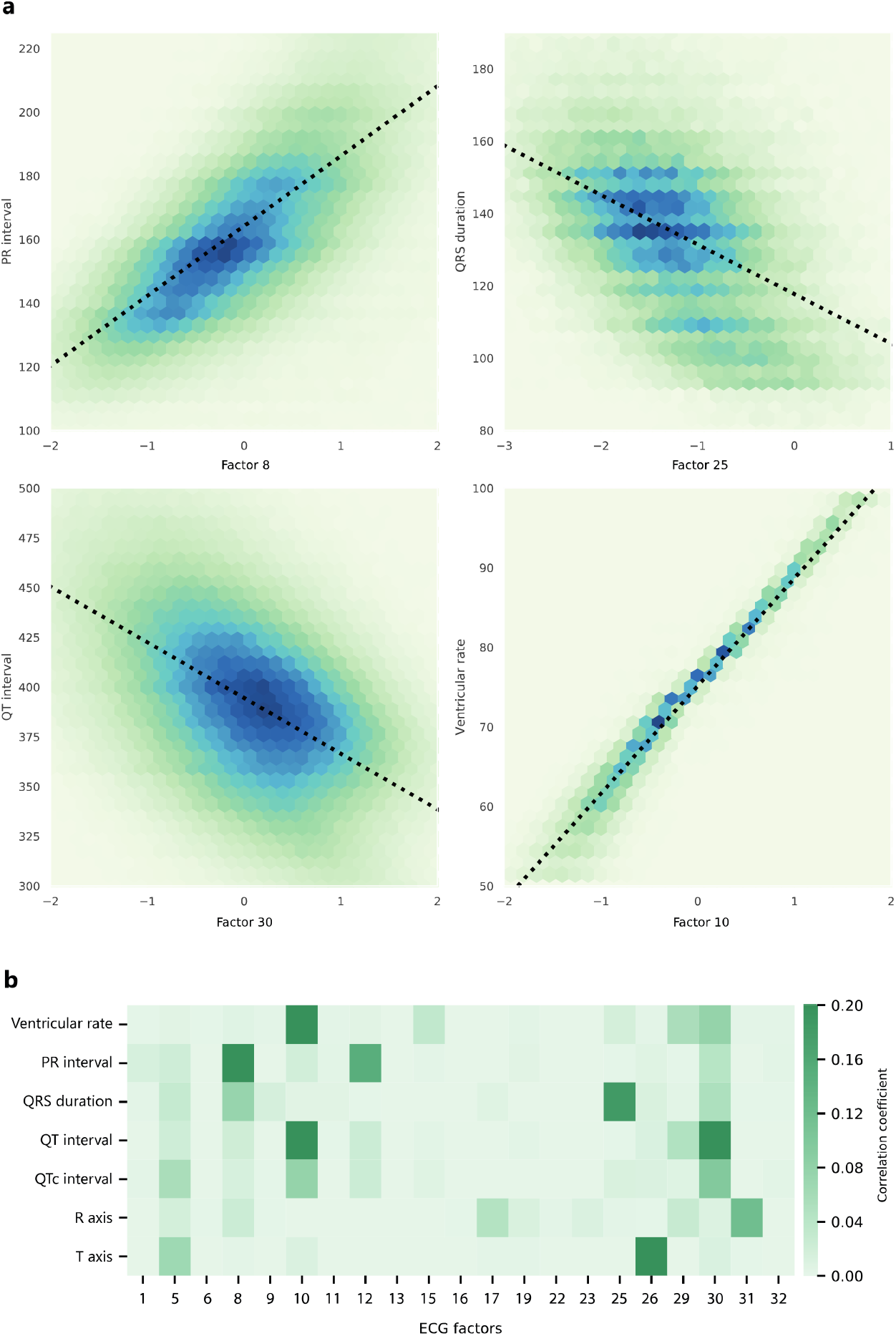
Relationship of the ECG factors with conventional ECG parameters. **a**. Hexagon plots where datapoints of ECG factor-ECG parameter pairs over all samples in the VAE dataset are binned into hexagon grids to relate values of factors 8, 25, 30, and 10 to the PR interval, QRS duration, QT interval and ventricular rate, respectively. **b**. Pearson correlation coefficients between ECG measures of ventricular rate, PR interval, QRS duration, QT interval, Bazett corrected QT interval, R-axis, and T-axis and ECG factor values over all samples in the VAE dataset.

### Performance and explainability for conventional ECG interpretation

The dataset for training the algorithms to perform conventional ECG interpretation consisted of 369,216 ECGs of 152,831 patients, while for validation the expert-annotated dataset was used, containing 965 ECGs (of 965 patients) of adequate quality. 343 (36%) of the ECGs had more than one diagnostic statement and sinus rhythm was the most prevalent (72%), while third degree AV block was the least prevalent (0.1%, Table 1). The mean AUROC of the explainable FactorECG pipeline was 0·94 [95% CI 0·92–0·96], compared to 0·73 [95% CI 0·65–0·81] for the rule-based MUSE algorithm and 0·96 [95% 0·94–0·98] for the ‘black box’ DNN. The explainable pipeline performed similarly for most diagnostic statements but was outperformed for diagnosis of left ventricular hypertrophy and low QRS voltage by the ‘black box’ DNN (Table 1). The conventional MUSE algorithm, that is currently used in clinical practice, performed worst for all diagnostic statements (Table 1). To understand which ECG factors were important for the pipeline to detect each ECG statement, we used the logistic regression’s coefficients as feature importance scores (Figure 4). The negative (blue) and positive (red) scores from Figure 4 can be related to the generated ECGs in the factor traversals after negative (blue) and positive (red) perturbations in Figure 2 and Supplementary Figure 2.

**Figure 4.**
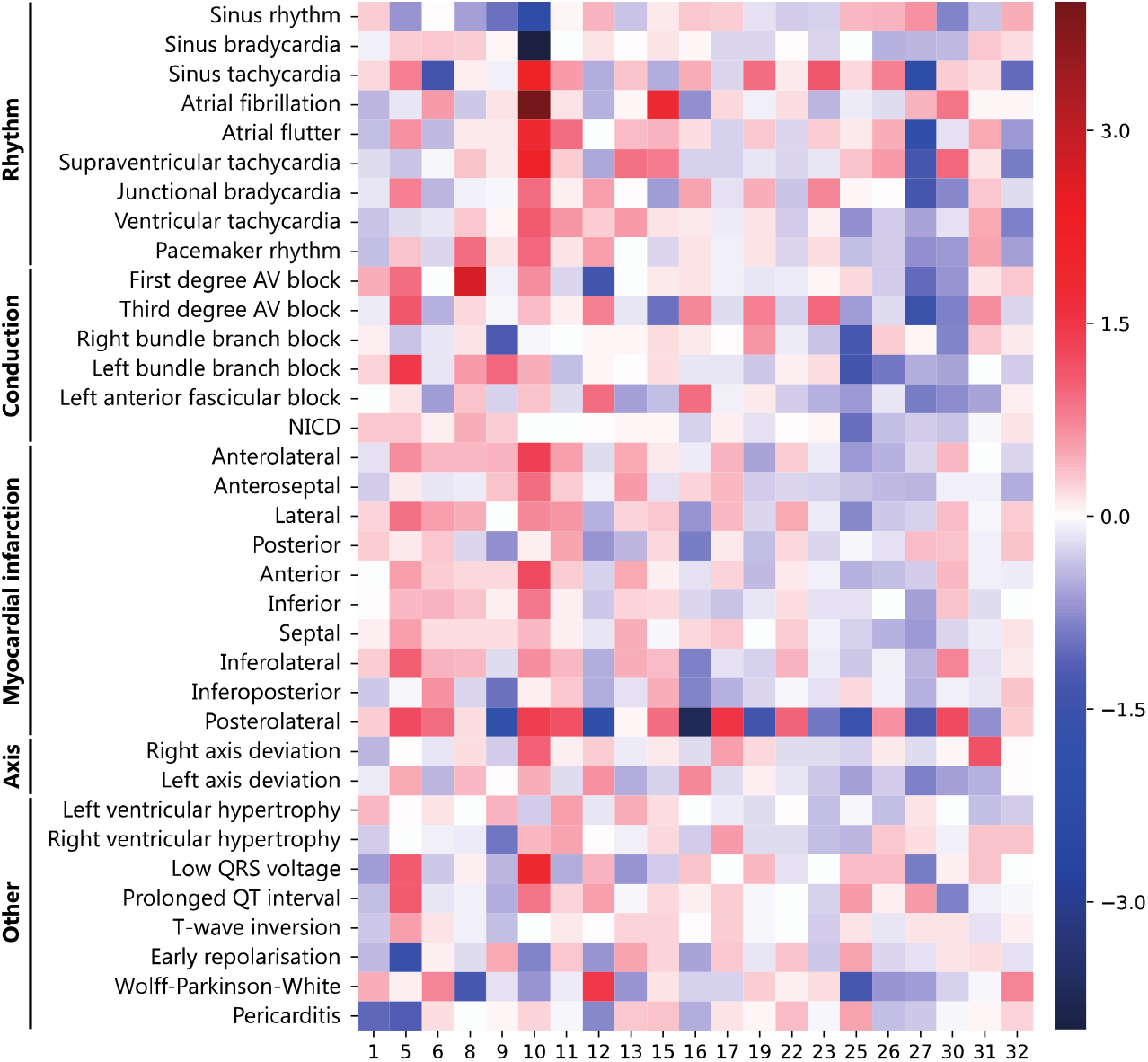
Importance score for each of the 32 factors in predicting 35 diagnostic ECG statements. Importance scores of each of the 32 factors in the logistic regression for all 35 diagnostic ECG statements are shown to relate which dimensions are important for diagnosis. High importance values indicate that a high value for the dimension is diagnostic for that abnormality, and vice versa. The negative (red) and positive (blue) scores can be related to the reconstructions after negative (red) and positive (blue) perturbations in Figure 2. Notably, factor 10 encodes ventricular frequency (as observed in Figures 2 and 3) and therefore has a high value in sinus tachycardia (red) and a low value in sinus bradycardia (blue). NICD: nonspecific intraventricular conduction delay.

### Performance and explainability for detection of reduced ejection fraction

For the algorithms to detect reduced ejection fraction, 39,603 matched ECG-echocardiogram pairs of 22,676 patients were available, of which 25% (5669 unique patients, first pair per patient used) was used for validation. 713 patients (13%) in the validation set had an ejection fraction below 35%. The explainable FactorECG pipeline achieved an AUROC of 0·90 (95% CI 0·89–0·91), in comparison to 0·91 (95% CI 0·90–0·93 for the ‘black box’ DNN. The most important model-level ECG factors for detecting reduced ejection fraction were high values in factors 5, 10 and 8 and low values in factors 25, 26, 1 and 30 (Supplementary Figure 3a). These correspond to negative T waves, higher ventricular rate, ST elevation, increased P-wave area and PR-interval, right bundle branch block, and left bundle branch block, respectively. Figure 5 shows a model- and individual patient-level explanation for the detection of reduced ejection fraction using the novel pipeline, in comparison to the post-hoc explainability methods used up until now.

**Figure 5.**
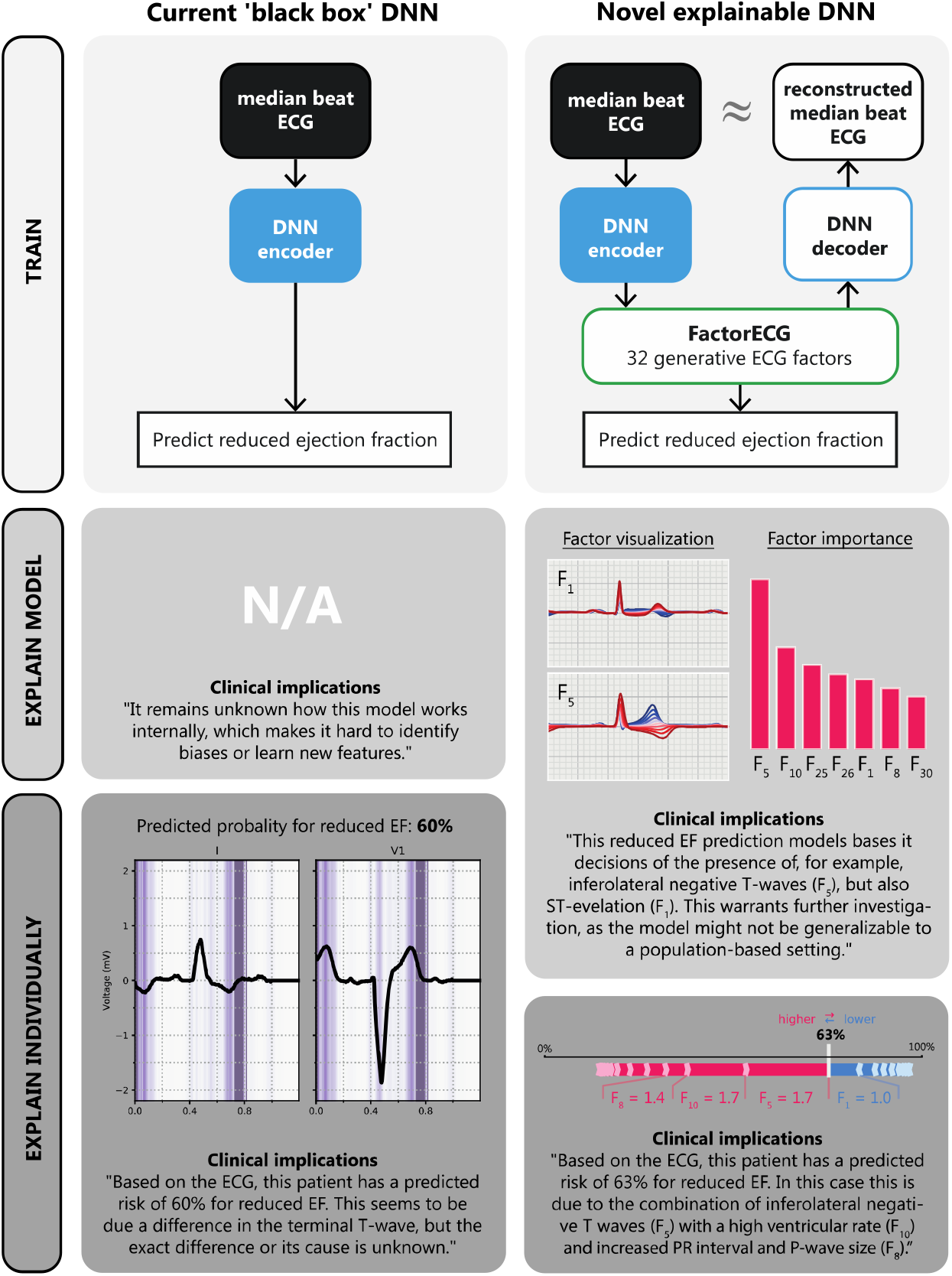
Comparison of architecture and model- and individual patient-level explainability using the novel inherently explainable approach as compared to post-hoc heatmap-based explainability for detection of reduced ejection fraction. The conventional ‘black box’ DNN contains only a single encoder to interpret the ECG. Afterwards, Guided Grad-CAM is applied to show what segments of the ECG were important for prediction on the patient-level. Model-level explainability is not possible. The novel explainable DNN (FactorECG) adds a generative part to the architecture, which allows for precise visualizations of the morphological ECG features. By combining factor SHAP importance scores and factor traversals, we obtain model-level explainability. Individual patient-level explainability is achieved using individual SHAP importance scores.

### Performance and explainability for prognosis of one-year mortality

For the models to predict one-year mortality, follow-up was available for 909,958 ECGs of 177,448 patients, of which 40% (70,979 unique patients, ECG sampled randomly per patient) was used for validation. 5334 patients (7.5%) in the validation set deceased within one year. The explainable FactorECG pipeline achieved an AUROC of 0·76 (95% CI 0·76–0·77), compared to 0·75 (95% CI 0·74– 0·76) for the ‘black box’ DNN. In contrast, an XGBoost model that included only age and sex had an AUROC of 0·65 (95% CI 0·64–0·66). The most important global ECG factors for prediction of one-year mortality were high values for factors 10, 5, 12 and 11, and low values for factors 1, 30, 9 and 27 (Supplementary Figure 3b). These correspond to an increased risk of one-year mortality with higher ventricular rate, inferolateral negative T-waves, ST-elevation, prolonged QT interval and anterior negative T-waves.

## Discussion

In this study, we demonstrate a novel pipeline that provides inherently explainable interpretation of ECGs, which consists of three major components: (i) a generative deep learning model that learned to summarize the underlying factors of variation of an ECG in 21 factors (the FactorECG), (ii) a visualization technique to provide insight into ECG morphology that these factors encode, and (iii) a common interpretable statistical method to perform diagnosis or prediction using the ECG factors (Figure 1). We investigated the FactorECG using visualizations and associations with conventional ECG parameters and diagnostic ECG statements to show that it encodes valid and relevant generative factors of ECG morphology. Moreover, when applying the novel explainable technique for conventional ECG interpretation and recently emerged use cases for the ECG, not only did it perform similarly to the ‘black box’ algorithms for these use cases, but it could also explain which morphological ECG features were important for prediction or diagnosis. This indicates that inherently explainable deep learning methods should be used to gain confidence in AI for clinical decision making, and more importantly, make it possible to identify biased or inaccurate models.

A longstanding assumption was that the high-dimensional and non-linear ‘black box’ nature of the currently applied DNNs was inevitable to gain the impressive performances shown by these algorithms.^5,13,22^ The major finding of the current study is that an inherently explainable DNN performs on par with the ‘black box’ algorithms in both conventional and novel tasks (Table 1), while also allowing for detection of biases or learning new features. Moreover, a main advantages of the current approach over previous attempts to open the ‘black box’ of DNNs using post-hoc explainability methods (i.e. heatmaps) is that the model is inherently explainable: we can reliably specify the morphology of the ECG feature, instead of only pointing at the location on the ECG’s time axis.^3,5,10,18^ Furthermore, our approach allows for both model-level (i.e. for the complete model by using factor traversals) and individual patient-level explanations (i.e. we know the values of the FactorECG for each patient’s ECG individually), while heatmap methods (such as Grad-CAM) are only usable for individual patient-level explanations.^19^ Moreover, recent studies have shown that saliency-based methods can be very unreliable in providing consequent annotation and can also show reassuring saliency maps when a model is completely untrained, stressing the need for better approaches to explain output of DNNs.^8,20^

We hypothesized that an ECG can be explained by a few underlying explanatory factors of variation and showed that it is possible to encode the median beat ECG morphology in 21 continuous factors, from which the ECG can be reconstructed with high precision. An online tool for clinicians to interactively visualize the factors can be found via https://decoder.ecgx.ai. When relating the ECG factor traversals (Figure 2 and Supplementary Figure 2) to diagnostic ECG statements and conventional ECG parameters (Figures 3 and 4), we were able to relate them to the underlying anatomical and (patho)physiological factors. For example, factor 10 has a clear linear relationship with ventricular frequency and therefore shows high values for sinus tachycardia and low values for sinus bradycardia. Moreover, the factor traversals (Figure 2) show the changes in the ECG associated to the ventricular frequency, such as the length of the QT interval and appearance of the T-wave of the previous beat. Factors 6, 23 and 27 account for the P-wave size and are related to diagnoses that involve the P-wave, such as junctional bradycardia and atrial fibrillation, while PR interval (or location of the P-wave) is encoded in factor 8. Factors 25, 26 and 30 encode ventricular conduction delays, such as right and left bundle branch block, while ventricular repolarization is mainly encoded in factors 1, 5, 9, 13 and 30. ST elevation is most prominent in factors 1 and 5, which are subsequently important for predicting diagnoses such as acute pericarditis and early repolarization. Next to these more common ECG variations, rare abnormalities are also represented, as for example Wolff-Parkinson-White pattern (with pre-excitation and short PR interval) is encoded using a combination of factors 8 and 12.

For the reduced ejection fraction model we found that the performance of the explainable FactorECG pipeline is equivalent to both the black box DNN in our dataset and in the original publication by Attia et al.^4^ Most important ECG indicators for reduced EF were consistent with previous findings that indicated similar features to be predictive of heart failure: inferolateral negative T-waves, increased ventricular rate, P-wave area, prolonged PR interval, RBBB, LBBB, but also inferolateral ST elevation.^21^ The importance of this latter feature illustrates that the DNN also picks up reduced ejection fraction due to acute ischemia. This could severely hamper the generalizability of such models for screening purposes in the general population as these patients are only present in large hospitals and is one of the reasons why explainable models are imperative.^8,22^ Hence, it may even be an explanation for the reduced performance in the previously published study where the ejection fraction model was externally validated in a population-based cohort.^23^

Although the model for one-year mortality performs worse than in the original paper by Raghunath et al., it does perform similarly to the ‘black box’ DNN on our dataset.^5^ The difference in performance is likely caused by differences in the population, as the predictive value of just age and sex is also lower than in the original paper. We observed that the predictors for one-year mortality are increasing age, higher ventricular frequency, negative T-waves and ST-depression and elevation and prolonged QT interval, which are all known risk factors for mortality.^24,25^

There are several limitations to acknowledge. Firstly, the algorithm is trained on a very large dataset with over 1 million ECGs, but we could not account for imbalance in ECG abnormalities due to the unsupervised nature of training. Therefore, less common ECG abnormalities might not be accurately encoded, as also demonstrated by the lower performance on for example ischemia classes and lower correlation coefficients of the reconstructed ECGs (Supplementary Table 1). Future studies could experiment with balancing the dataset based on labelled abnormalities and the effect it may have on encoding rare ECG abnormalities. Secondly, the reduced performance of the explainable model in diagnosing low QRS voltage and left ventricular hypertrophy is most likely due to the inability of the VAE to always reconstruct the amplitude of the R-wave correctly (Supplementary Table 1). Further research in the field of generative models for ECGs is needed to address this limitation and to improve the reconstruction quality.

Future studies should focus on evaluating the use of inherently explainable deep learning methods on other ECG tasks, as the dimensionality reduction of our algorithm to 21 factors broadens the usability of DNNs greatly to much smaller labeled datasets than before. Another important perspective is using the approach on full 10-second rhythm ECGs, to take additional ECG information into account. Rhythm disorders that are not visible in the median ECG beat, such as second-degree AV block and premature ventricular and atrial complexes, could add interesting information to the model.

In conclusion, we leveraged a large dataset of over 1 million ECGs to train a generative DNN that learned 21 valid underlying anatomical and (patho)physiological explanatory factors of variation in median beat 12-lead ECG data. We showed that our pipeline is not only able to interpret ECGs with highly accurate performance on par with ‘black box’ DNNs but can also explain which ECG morphologies were important. These findings demonstrate that inherently explainable ECG models should be the future of ECG interpretation, as they allow reliable clinical interpretation of these models without performance reduction, while also broadening their applicability to many other (rare) diseases.

## Supporting information

Supplementary Animation

## Data Availability

The training datasets used in this study are not openly available due to privacy concerns. The expert-annotated test set is available upon request to the corresponding author. The decoder for the FactorECG is publicly available at https://decoder.ecgx.ai. The code for training and evaluating the β-VAE and the black box DNN is available upon request to the corresponding author.

## Data availability

The training datasets used in this study are not openly available due to privacy concerns. The expert-annotated test set is available upon request to the corresponding author.

## Code availability

The decoder for the FactorECG is publicly available at https://decoder.ecgx.ai. The code for training and evaluating the β-VAE and the black box DNN is available upon request to the corresponding author.

## Acknowledgments

We gratefully acknowledge the support of NVIDIA Corporation with the donation of the Titan Xp GPU used for this research.

## Ethics committee approval

This study was approved by the University Medical Center Utrecht ethical committee with number 18-827.

## Author contributions

Conceptualization: R.R.v.d.L., M.N.B., D.K.G. and R.v.E.; Data curation: R.R.v.d.L., M.N.B. and R.v.E.; Formal analysis: R.R.v.d.L., M.N.B. and D.K.G.; Funding acquisition: R.R.v.d.L., R.J.H. and P.A.D.; Investigation: R.R.v.d.L. and M.N.B.; Methodology: R.R.v.d.L. and M.N.B.; Supervision: R.J.H., P.A.D., D.K.G. and R.v.E.; Writing – original draft: R.R.v.d.L. and M.N.B.; Writing - review & editing: K.T., A.S., S.v.D., P.D.L., R.J.H., P.v.d.H., P.A.D., D.K.G. and R.v.E.

## References

1 Leur RR van de, Boonstra MJ, Bagheri A, et al. Big Data and Artificial Intelligence: Opportunities and Threats in Electrophysiology. Arrhythmia Electrophysiol Rev 2020; 9: 146–54.

2 Hannun AY, Rajpurkar P, Haghpanahi M, et al. Cardiologist-level arrhythmia detection and classification in ambulatory electrocardiograms using a deep neural network. Nat Med 2019; 25: 65– 9.

3 Leur RR van de, Blom LJ, Gavves E, et al. Automatic Triage of 12-Lead Electrocardiograms Using Deep Convolutional Neural Networks. J Am Heart Assoc 2020; 9. DOI:10.1161/jaha.119.015138.

4 Attia ZI, Kapa S, Lopez-Jimenez F, et al. Screening for cardiac contractile dysfunction using an artificial intelligence–enabled electrocardiogram. Nat Med 2019; 25: 70--74.

5 Raghunath S, Cerna AEU, Jing L, et al. Prediction of mortality from 12-lead electrocardiogram voltage data using a deep neural network. Nat Med 2020; 26: 886--891.

6 Ghassemi M, Oakden-Rayner L, Beam AL. The false hope of current approaches to explainable artificial intelligence in health care. Lancet Digital Heal 2021; 3: e745–50.

7 Kundu S. AI in medicine must be explainable. Nat Med 2021; : 1–1.

8 Rudin C. Stop explaining black box machine learning models for high stakes decisions and use interpretable models instead. Nat Mach Intell 2019; 1: 206–15.

9 Goodman B, Flaxman S. European Union Regulations on Algorithmic Decision-Making and a “Right to Explanation.” Ai Mag 2017; 38: 50–7.

10 Leur RR van de, Taha K, Bos MN, et al. Discovering and Visualizing Disease-Specific Electrocardiogram Features Using Deep Learning: Proof-of-Concept in Phospholamban Gene Mutation Carriers. Circulation Arrhythmia Electrophysiol 2021; 14. DOI:10.1161/circep.120.009056.

11 Kwon J, Cho Y, Jeon K-H, et al. A deep learning algorithm to detect anaemia with ECGs: a retrospective, multicentre study. Lancet Digital Heal 2020; 2: e358–67.

12 Kingma DP, Welling M. Auto-Encoding Variational Bayes. In: Bengio Y, LeCun Yann, eds. 2nd International Conference on Learning Representations. Banff, AB, Canada: Conference Track Proceedings, 2014. http://arxiv.org/abs/1312.6114.

13 Higgins I, Matthey L, Pal A, et al. beta-VAE: Learning Basic Visual Concepts with a Constrained Variational Framework. In: 5th International Conference on Learning Representations. Toulon, France: Conference Track Proceedings, 2017.

14 Bos MN, Leur RR van de, Vranken JF, et al. Automated Comprehensive Interpretation of 12-lead Electrocardiograms Using Pre-trained Exponentially Dilated Causal Convolutional Neural Networks. 2020 Comput Cardiol 2020; 00: 1–4.

15 Chen T, Guestrin C. XGBoost: A Scalable Tree Boosting System. In: Proceedings of the 22nd ACM SIGKDD International Conference on Knowledge Discovery and Data Mining. East Lansing, MI, USA: ACM, 2016: 785–94.

16 Lundberg SM, Lee S-I. A Unified Approach to Interpreting Model Predictions. In: Guyon I, Luxburg U.V., Bengio S., et al., eds. Advances in Neural Information Processing Systems 30. Curran Associates, Inc., 2017: 4765--4774.

17 Collins GS, Reitsma JB, Altman DG, Moons KGM. Transparent Reporting of a multivariable prediction model for Individual Prognosis Or Diagnosis (TRIPOD): The TRIPOD Statement. Ann Intern Med 2015; 162: 55.

18 Kwon J, Lee SY, Jeon K, et al. Deep Learning–Based Algorithm for Detecting Aortic Stenosis Using Electrocardiography. J Am Heart Assoc 2020; 9: e014717.

19 Selvaraju RR, Cogswell M, Das A, Vedantam R, Parikh D, Batra D. Grad-CAM: Visual Explanations from Deep Networks via Gradient-Based Localization. 2017 Ieee Int Conf Comput Vis Iccv 2017; 2017-Octob: 618--626.

20 Adebayo J, Gilmer J, Muelly M, Goodfellow I, Hardt M, Kim B. Sanity Checks for Saliency Maps. Advances in Neural Information Processing Systems 2018; 2018-Decem: 9505--9515.

21 O’Neal WT, Mazur M, Bertoni AG, et al. Electrocardiographic Predictors of Heart Failure With Reduced Versus Preserved Ejection Fraction: The Multi-Ethnic Study of Atherosclerosis. J Am Heart Assoc 2017; 6. DOI:10.1161/jaha.117.006023.

22 Yao X, McCoy RG, Friedman PA, et al. ECG AI-Guided Screening for Low Ejection Fraction (EAGLE): Rationale and design of a pragmatic cluster randomized trial. Am Heart J 2020; 219: 31–6.

23 Attia IZ, Tseng AS, Benavente ED, et al. External validation of a deep learning electrocardiogram algorithm to detect ventricular dysfunction. International Journal of Cardiology 2021. DOI:10.1016/j.ijcard.2020.12.065.

24 Kannel WB, Kannel C, Paffenbarger RS, Cupples LA. Heart rate and cardiovascular mortality: The Framingham study. Am Heart J 1987; 113: 1489–94.

25 Porthan K, Viitasalo M, Jula A, et al. Predictive value of electrocardiographic QT interval and T-wave morphology parameters for all-cause and cardiovascular mortality in a general population sample. Heart Rhythm 2009; 6: 1202–1208.e1.

