## Supplementary Material for "Inherently explainable deep neural network-based interpretation of electrocardiograms using variational auto-encoders"

**Supplementary Methods**

*Data acquisition for training and validation of the VAE*

ECGs were recorded using a General Electric MAC V, 5000 or 5500 device and acquired at either 250 or 500 Hz. Linear interpolation was used to resample all recordings to 500 Hz. The 10-second ECGs were transformed into 1.2-second median beats by aligning all QRS-complexes of the same morphology (e.g., excluding premature ventricular complexes) and calculating the median voltage to generate a representative P-QRS-T complex.

*Data acquisition for training and validation of the algorithms to detect conventional diagnostic ECG statements*

For training of the algorithms to detect conventional diagnostic ECG statements, the free text ECG annotations were converted into standardized ECG statements (e.g. sinus rhythm, left bundle branch block) according to the American Heart Association’s Electrocardiography Diagnostic Statement List using a text mining-based approach described before.^1,2^ For comparison, the statements as provided by the built-in MUSE 12SL algorithm were also exported. A single recording can have multiple diagnostic statements.

The following 35 diagnostic statements were included: sinus rhythm, sinus bradycardia, atrial fibrillation, sinus tachycardia, atrial flutter, supraventricular tachycardia, junctional bradycardia, ventricular tachycardia, pacemaker rhythm, right axis deviation, left axis deviation, left ventricular hypertrophy, right ventricular hypertrophy, first-degree AV block, third-degree AV block, right bundle branch block, left bundle branch block, left anterior fascicular block, nonspecific intraventricular conduction delay, myocardial infarction (anterolateral, anteroseptal, lateral, posterior, anterior, inferior, infarct, inferolateral, inferoposterior and posterolateral), low QRS voltage, prolonged QT interval, T-wave inversion, early repolarization, Wolff-Parkinson-White pattern and acute pericarditis. Rhythm disorders that are not present in the median beat, such as premature ventricular complexes or second-degree AV block, were not evaluated. The ECGs of 80% of the patients were included in the training set, while the ECGs of the rest of the patients (20%) were used in a holdout validation set, while ensuring that all patients that are included in the test dataset were removed from the training dataset.

For the expert-annotated validation set, all ECGs were interpreted independently by two cardiologists, that were blinded to the other interpretation. In case of disagreement in one of the statements, a third cardiologist was consulted, and the majority vote was used to determine the final statement. All recordings that were deemed technically insufficient by the cardiologists were excluded.

*Data acquisition and annotation for training and testing of reduced ejection fraction and one year mortality models*

Firstly, for the prediction of one-year mortality using the ECG, we selected patients using the same approach as Raghunath et al.^3^ From the full ECG database, we selected all patients with at least 1 year of follow-up available, or death within 1 year, from the acquisition date of the ECG onwards (n = 909,958). Age and sex for these patients were derived from the electronic health record and mortality data was retrieved from the nationwide Personal Records Database. The dataset was split into training and test in a 60:40 manner on the patient level and for the test dataset one random ECG per patient was sampled. This was done to avoid over-representation of sicker patients with more ECGs and is considered the most representative strategy when applied in practice.

Secondly, for the diagnosis of patients with reduced ejection fraction (EF), we applied a similar strategy to the paper by Attia et al.^4^ We retrieved all echocardiograms (n = 57,965) from the digital database between April 1^st^ 2008 and October 31th 2019 and matched those to the closest ECG, acquired within a 30 day timeframe. This resulted in 39,603 matched pairs, and the EF was extracted from the echocardiogram report using a standard hierarchical sequence: first 3-D based approximation, then a biplane approach, a 2-D method or M-mode method. The EF was dichotomized at a 35% cut-off. A 75:25 train:test split was made on the patient level and in the test set only the first ECG-echocardiogram pair was used for analysis.

*VAE model architecture and training*

The overall architecture of the β-VAE is shown in Supplemental Figure 1, where the encoder $f_{\phi}\left( \boldsymbol{x} \right)$and decoder $f_{\theta}\left( \boldsymbol{z} \right)$ are convolutional neural networks specifically designed for the temporal nature of ECGs and the remaining components are as in standard VAEs.^5^ The β-VAE loss function is used to employ a higher weight on the Kullback-Leibler divergence (KLD) term in the objective to enforce disentanglement. By increasing the β-value in β-VAE models, the model is increasingly enforced to encode 12-lead ECG data in to lower-dimensional latent spaces of independent standard normal distributions. The prior over the latent variables is chosen to be the centered isotropic multivariate Gaussian $p_{\boldsymbol{\theta}}\left( \mathbf{z} \right)\mathcal{=N}\left( \mathbf{z};\mathbf{0},\mathbf{I} \right)$. We let both approximate posteriors $p_{\theta}\left( \mathbf{x} | \mathbf{z} \right)$ and $q_{\phi}\left( \mathbf{z} | \mathbf{x} \right)$ to be multivariate Gaussians, where the mean and standard deviations of $\mathbf{x}$ and $\mathbf{z}$ posterior are parameterized by the outputs of the $f_{\theta}\left( \boldsymbol{z} \right)$ and $f_{\phi}\left( \mathbf{x} \right)$ layers, respectively. The linear layers for learning the standard deviations use the SoftPlus activation function plus a non-negative small number $\epsilon$, for constraining the standard deviations to be non-negative and to avoid numerical issues that might occur when computing the likelihoods of Gaussian variables if the value becomes very close to zero.

For the encoder, we choose a convolutional deep neural network with an architecture of exponentially dilated causal convolutions to encode median beat ECG data. This network structure has shown to work well with median beat ECG data and is inspired by Van den Oord et al and Bos et al.^6–8^ The encoder structure $f_{\phi}\left( \boldsymbol{x} \right)$is illustrated in Supplemental Figure 1. The encoder is first composed of eight 1-dimensional causal convolution blocks to transform the 12x600-sized median beat ECG data to 64 600-dimensional feature maps.^9^ Subsequently, we employ a 1-dimensional adaptive max pooling layer to squeeze the temporal dimension resulting in a 64-dimensional representation. This is followed by two parallel 64-to-32 linear layers to transform the representation to mean and standard deviation parameters for the Gaussian distribution in the latent space, where we apply a SoftPlus activation function plus a non-negative small number $\epsilon$ of 0.001 on the standard deviation linear layer. Each causal convolution block consists of a combination of causal convolutions, weight normalizations, leaky ReLUs and residual connections. The causal convolution is a result of first applying a convolution and thereafter truncating the output, and the residual connection is only used when upsampling or downsampling the number of input channels. The dilation parameter used in the causal convolutional layer is doubled in each subsequent causal convolution block from 1 to 128. The first convolutional layer transforms the 12 input channels to 128 output channels, and thereafter the number of channels is kept constant at 128. The last causal convolution block downsamples the number of channels from 128 to 64. The causal convolutions used a kernel size of 5. As a result of employing causal convolutions and subsequently adaptive max pooling to extract features from the temporal dimensions, the length of the input channels is variable, though we use a length of 600 for median beat data.

For the decoder, we propose to use a mirrored version of the encoder to reconstruct lower-dimensional representations of the original electrocardiograms with continuous outputs as illustrated in Figure 1. To mirror the max pooling layer, the 32-dimensional representation $z$ is first transformed to a 64-dimensional vector using a 32-to-64 linear layer. Then, the 64-dimensional vector is transformed to a 64x600-sized matrix by using a 64-to-$\left( 64\cdot600 \right)$ linear layer and reshaping the output to 64x600. Subsequently, this is deconvolved in a mirrored version of the stack of exponentially dilated causal convolution blocks, effectively reconstructing high-level ECG features to the original median beat ECG. The deconvolution is performed by replacing the 1-dimensional convolution layers by 1-dimensional transposed convolution layers. The dilation parameter used in the causal convolutional layer starts at 128 and is halved each subsequent causal convolution block, from 128 to 1. Finally, we flatten the output of the mirrored stack of causal convolution blocks from size 12x600 to a $\left( 12\cdot600 \right)$-dimensional vector and use two parallel $\left( 12\cdot600 \right)$-to-$\left( 12\cdot600 \right)$ linear layers to learn the mean and standard deviation parameters for the Gaussian distribution of each point in the reconstructed 12x600 ECG, where we apply a SoftPlus activation function plus a non-negative small number $\epsilon$ of 0.001 on the standard deviation linear layer. The final reconstruction is obtained by reshaping $\mu_{z}$ to 12x600.

The two most important hyperparameters in the β-VAE were the number of ECG factors and the β

-value. For both, values of 8, 16, 32, 64 and 128 were evaluated. Considering that increasing the β-term results in higher reconstruction errors, we chose a β that resulted in a good trade-off for good reconstruction with adequate disentanglement in significant factors, which was assessed using the factor traversals and the reconstruction error. Moreover, increasing the number of ECG factors above 32 did not yield an increase in significantly contributing factors (i.e., factors that encode variation), therefore this value was selected. The VAE model was trained on the entire VAE train set, using the Adam optimizer with a learning rate of 0.001, and batch size was set at 128.^15^ Models were trained for 200 epochs, and we selected a final model that yielded the lowest loss on the evaluation set. To evaluate the training progress of the VAE and to determine which epoch to select as the inference model, 10% of this data was left out of training.

The VAE architecture and model training processes were implemented using PyTorch (version 1.7.0+cu110) in Python (version 3.6.7).^10^ All training was performed using an NVIDIA Titan Xp GPU, with an average duration of ~45 minutes per epoch.

*Baseline ‘black box’ DNN architecture*

We developed baseline ‘black box’ DNNs for the ECG interpretation, reduced ejection fraction and one-year mortality tasks that used the median beat 12-lead ECG data as input. The DNN is constructed with the same architecture as the encoder part of the β-VAE. However, the last two parallel 64-to-32 linear layers are replaced by one 64-to-*n* linear layer to transform the 64-dimensional representation resulting from the adaptive max pooling layer to a network output equal to the number of detectable classes *n*. For the training criterion we used a binary focal loss function with a gamma of 2 on the output scores. The output predictions were obtained by applying a sigmoid function on the output scores.

*Explainable DNN pipeline training for ECG interpretation*

As it provided maximum interpretability, we trained binary logistic regression models for each of the 35 diagnostic ECG statements on the FactorECGs with 32 continuous factors. As a preprocessing step, each ECG in the logistic regression training set was encoded into its representative 32 ECG factors using the β-VAE model, and data for each factor was standardized by removing the mean and scaling to unit variance. We used the Limited-memory Broyden–Fletcher–Goldfarb–Shanno solver, L2 penalty, an inverse of regularization strength value of 1·0, a tolerance for stopping criteria of 1x10^-4^, and a maximum number of iterations taken for the solvers to converge of 100. To compensate for the imbalance in proportion of negative/positive samples for each class, we used grid search on the class weight to determine a value for each model from 1 to 100. Repeated stratified k-fold cross validation was employed to find the best estimator using 10 splits, 3 repeats, and compared based on the geometric mean of the specificity and sensitivity. The importance of the factors was assessed using the coefficient value of the logistic regression estimator.

*Explainable DNN pipeline training for reduced ejection fraction and one-year mortality*

For the secondary analysis, as the aim was maximum performance, we trained two extreme gradient boosting decision trees (XGBoost) for the mortality and ejection fraction task using the 32 ECG factors.^11^ The following hyperparameters were optimized using Bayesian optimization: maximum tree depth, learning rate, gamma, minimum child weight, overall subsampling ratio, subsampling ratio per tree and number of estimators.^12^ Hyperparameters were evaluated on the training dataset using 10-fold cross-validation, aiming for the highest area under the receiving operating curve (AUROC). The model with the best hyperparameters was subsequently trained on the complete training set and tested on the test set.

**References**

1 Leur RR van de, Blom LJ, Gavves E, *et al.* Automatic Triage of 12-Lead Electrocardiograms Using Deep Convolutional Neural Networks. *J Am Heart Assoc* 2020; 9. DOI:10.1161/jaha.119.015138.

2 Mason JW, Hancock EW, Gettes LS. Recommendations for the Standardization and Interpretation of the Electrocardiogram. *Circulation* 2007; 115: 1325–32.

3 Raghunath S, Cerna AEU, Jing L, *et al.* Prediction of mortality from 12-lead electrocardiogram voltage data using a deep neural network. *Nat Med* 2020; 26: 886--891.

4 Attia ZI, Kapa S, Lopez-Jimenez F, *et al.* Screening for cardiac contractile dysfunction using an artificial intelligence–enabled electrocardiogram. *Nat Med* 2019; 25: 70--74.

5 Kingma DP, Welling M. Auto-Encoding Variational Bayes. In: Bengio Y, LeCun Yann, eds. 2nd International Conference on Learning Representations. Banff, AB, Canada: Conference Track Proceedings, 2014. http://arxiv.org/abs/1312.6114.

6 Oord A van den, Dieleman S, Zen H, *et al.* WaveNet: A Generative Model for Raw Audio. *Neural Comput* 2016; 21: 793--830.

7 Bos MN, Leur RR van de, Vranken JF, *et al.* Automated Comprehensive Interpretation of 12-lead Electrocardiograms Using Pre-trained Exponentially Dilated Causal Convolutional Neural Networks. *2020 Comput Cardiol* 2020; 00: 1–4.

8 Leur RR van de, Taha K, Bos MN, *et al.* Discovering and Visualizing Disease-Specific Electrocardiogram Features Using Deep Learning: Proof-of-Concept in Phospholamban Gene Mutation Carriers. *Circulation Arrhythmia Electrophysiol* 2021; 14. DOI:10.1161/circep.120.009056.

9 Franceschi J-Y, Dieuleveut A, Jaggi M. Unsupervised Scalable Representation Learning for Multivariate Time Series. In: Wallach H, Larochelle H, Beygelzimer A, Alche-Buc F d\textquotesingle, Fox E, Garnett R, eds. Advances in Neural Information Processing Systems. Curran Associates, Inc., 2019: 4650--4661.

10 Paszke A, Gross S, Massa F, *et al.* PyTorch: An Imperative Style, High-Performance Deep Learning Library. In: Advances in Neural Information Processing Systems. 2019. http://papers.neurips.cc/paper/9015-pytorch-an-imperative-style-high-performance-deep-learning-library.pdf.

11 Chen T, Guestrin C. XGBoost: A Scalable Tree Boosting System. In: Proceedings of the 22nd ACM SIGKDD International Conference on Knowledge Discovery and Data Mining. East Lansing, MI, USA: ACM, 2016: 785–94.

12 Snoek J, Larochelle H, Adams RP. Practical Bayesian Optimization of Machine Learning Algorithms. In: Pereira F, Burges C J C, Bottou L, Weinberger K Q, eds. Advances in Neural Information Processing Systems 25. Curran Associates, Inc., 2012: 2951--2959.

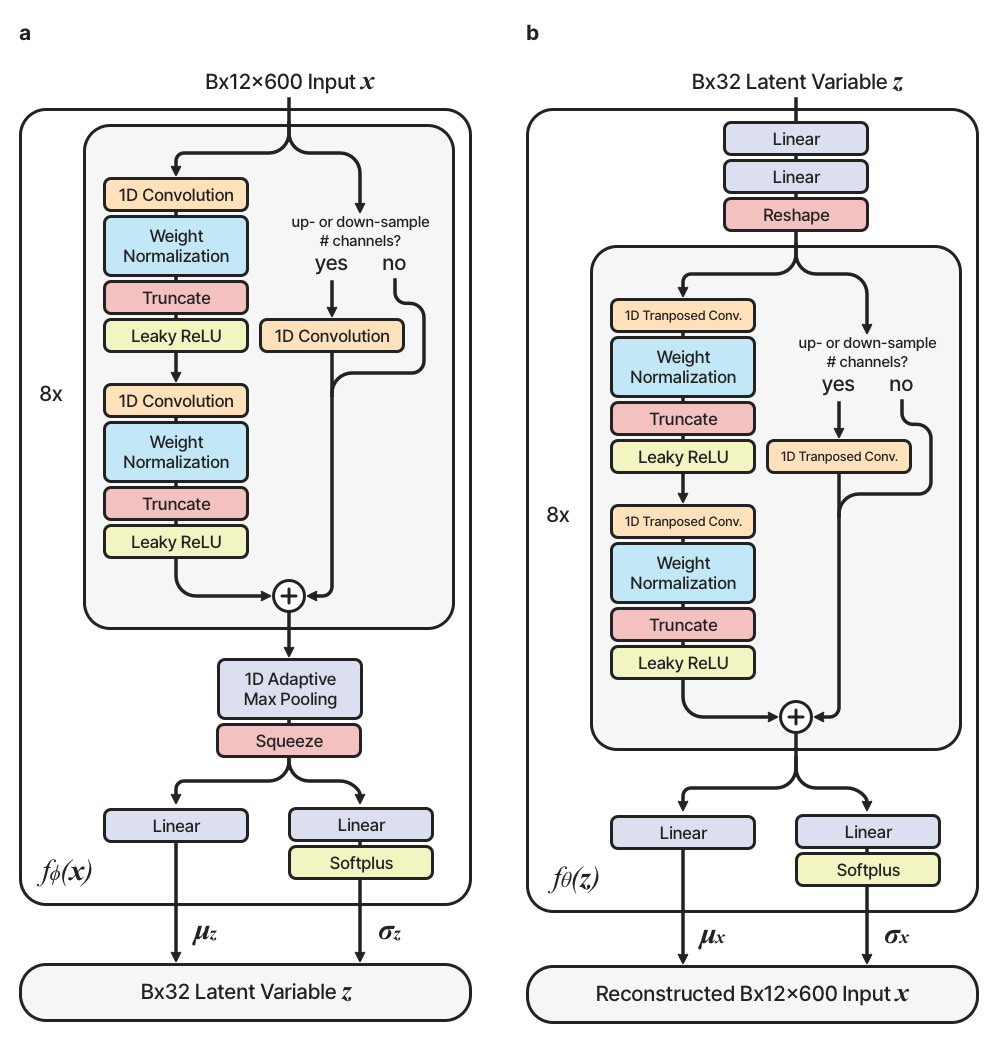
**Supplementary Figure 1. | The encoder and decoder part of the β-VAE architecture**

**(a)** Schematic diagrams of the variational network, the encoder part, that comprises the variational autoencoder architecture. The variational net is used to encode B × 12 × L-sized ECG data x into a lower-dimensional latent variable z. **(b)** Schematic diagram of the generative network, the decoder part, that comprises the variational autoencoder architecture. The generative network is used to reconstruct the original ECG data x from z. B and L stand for batch size, and temporal length of the ECGs, respectively.

*See separately attached pdf.*

**Supplementary Figure 2.** Factor traversals for all 21 ECG factors that hold significant information for correctly reconstructing ECG data. Each row corresponds to the factor traversal for one ECG factor and the columns to a subset of the 12 leads. The factor traversal for one row is obtained by starting with a ‘mean’ FactorECG where all factors are zero and adding offsets for that factor in a range of -5 to 5. The generated ECGs are then plotted where red represents high values for that factor and blue low values.

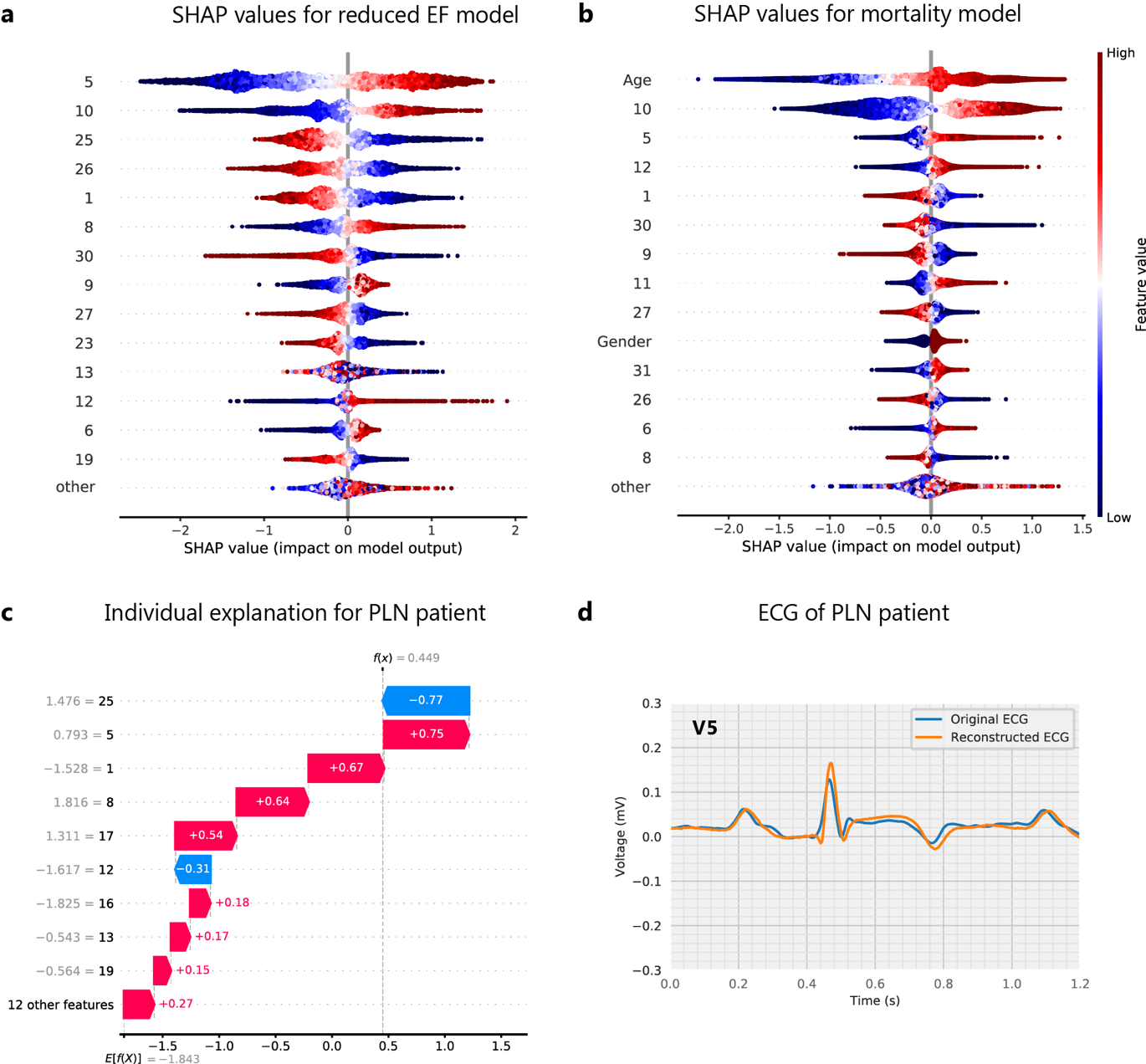

**Supplemental Figure 3. Explanations for the one-year mortality and reduced EF models using SHAP values.**

**a.** The most important global ECG factors for detecting reduced ejection fraction computed using SHAP values. Importance is ordered from top-to-bottom and coloring corresponds to the reconstructed ECGs in Figure 2. **b.** The most important global ECG factors for predicting one-year mortality. Importance is ordered from top-to-bottom and coloring corresponds to the reconstructed ECGs in Figure 2.

| Diagnostic ECG statement | Mean correlation coefficient |
| --- | --- |
| Acute pericarditis | 0.92 |
| Early repolarisation | 0.92 |
| Sinus rhythm | 0.91 |
| Sinus bradycardia | 0.91 |
| First degree AV block | 0.89 |
| Left bundle branch block | 0.89 |
| Third degree AV block | 0.89 |
| Left ventricular hypertrophy | 0.88 |
| Sinus tachycardia | 0.88 |
| Atrial fibrillation | 0.87 |
| Prolonged QT interval | 0.86 |
| Anteroseptal infarction | 0.85 |
| Supraventricular tachycardia | 0.84 |
| Atrial flutter | 0.84 |
| Left anterior fascicular bloack | 0.83 |
| Junctional bradycardia | 0.83 |
| T-wave inversion | 0.82 |
| Inferolateral infarction | 0.82 |
| Left axis deviation | 0.82 |
| Nonspecific intraventricular conduction delay | 0.81 |
| Right ventricular hypertrophy | 0.8 |
| Pacemaker rhythm | 0.79 |
| Right bundle branch block | 0.79 |
| Right axis deviation | 0.78 |
| Low QRS voltage | 0.77 |
| Inferoposterior infarction | 0.7 |
| Ventricular tachycardia | 0.7 |
| Lateral infarction | 0.67 |
| Anterior infarction | 0.65 |
| Inferior infarction | 0.62 |

**Supplementary Table 1.** Mean Pearson correlation coefficients between the original and reconstructed ECGs by the VAE for all 35 diagnostic ECG statements separately.
