## Supplementary figures and images for "Inherently explainable deep neural network-based interpretation of electrocardiograms using variational auto-encoders"

### Supplementary Figure 2

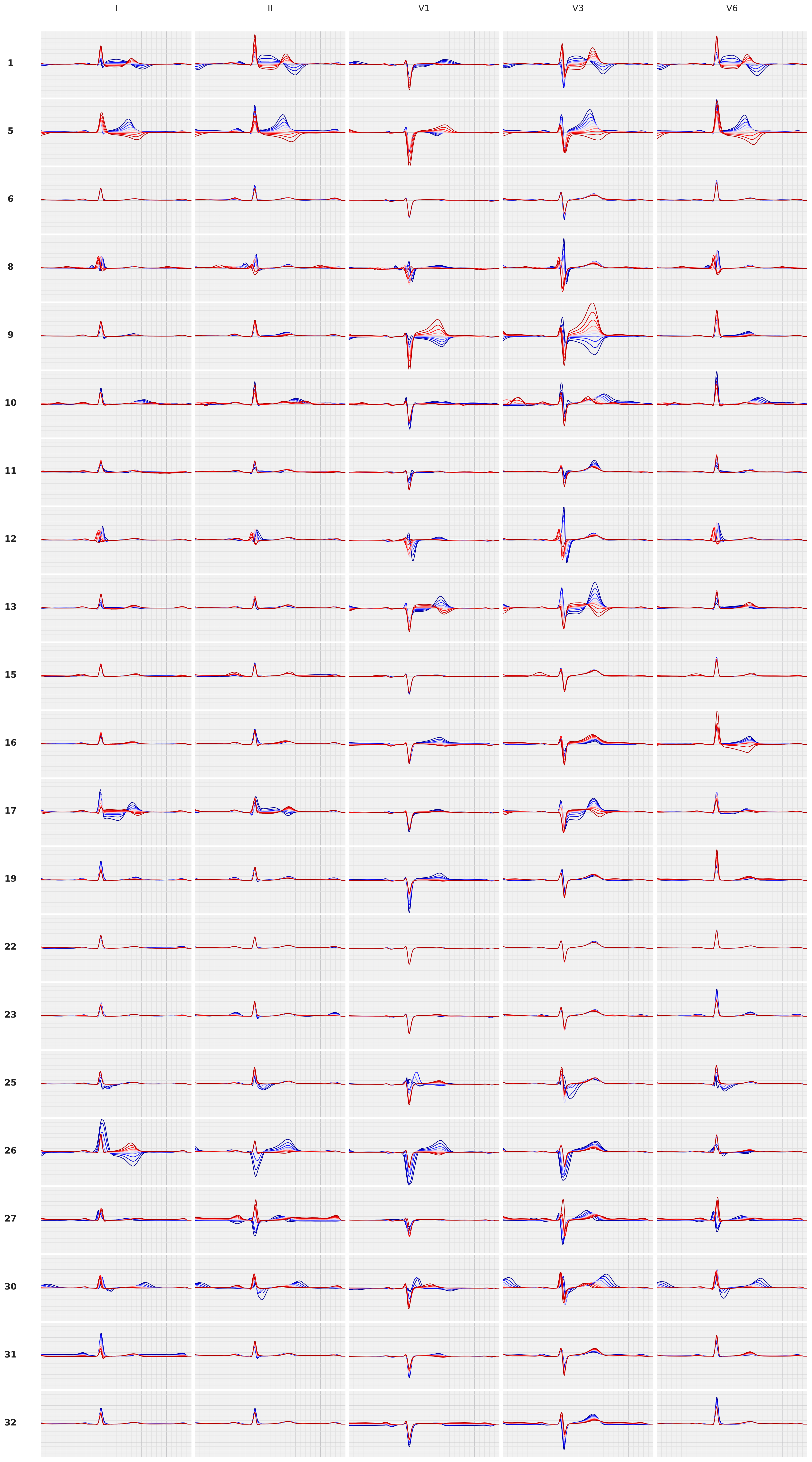
